# Impact of Life’s Essential Eight and Inflammatory Markers on Long-term Cardiovascular Disease Risk and All-cause Mortality: The Heart SCORE Study

**DOI:** 10.1101/2025.06.02.25328831

**Authors:** Claudia E. Bambs, Ian Pollack, Justin Swanson, Jiaxuan Duan, Christopher McKennan, Kevin Kip, Daniel Buysse, Steven E. Reis, Anum Saeed

## Abstract

**Background:** Life’s Essential 8 (LE8) are the key health behaviors and factors for improving and maintaining cardiovascular health, as defined by the American Heart Association. However, despite the association of cardiovascular diseases with inflammation, LE8 does not include inflammatory markers as a component. We analyzed the longitudinal association of LE8 components and inflammatory measures with health outcomes among a community-based population.

**Methods:** Baseline LE8 metrics and inflammatory markers (interleukin-6 [IL-6] and high sensitivity C-reactive protein [hsCRP]) were measured among 1,869 participants (age 59±7.5 years, 41.9% Black) in the longitudinal Heart SCORE study. Cox-proportional hazard ratios were used to assess the association of incident atherosclerotic cardiovascular disease (ASCVD) and all-cause mortality across quartiles of LE8 score, and to assess inflammatory markers as an independent risk factor.

**Results:** Compared to the lowest quartile, all higher quartiles of LE8 Score were significantly associated with lower risk of all-cause mortality and ASCVD over a median of 12 years of follow-up. After adjusting for all other LE8 metrics as well as by age, sex, race and socioeconomic risk, ideal level of blood glucose was the single most significant factor associated with lower risk of ASCVD (HR 0.26 [0.12-0.58], p=0.001). For all-cause mortality, no smoking emerged as the main protective LE8 component (HR 0.29 [0.16-0.53], p<0.001). When added to LE8, interleukin-6 (IL-6) was independently associated with ASCVD (HR 1.54 [1.05-2.25], p=0.03),while lower IL-6 was associated with lower all-cause mortality (HR 0.52 [0.30-0.90] p=0.02). HsCRP was not associated with either outcome.

**Conclusions:** Higher LE8 Score and metrics are associated with lower risk of all-cause and ASCVD mortality as well as non-fatal events in this longitudinal cohort of White and Black adults. Additionally, elevated IL-6 levels were associated with higher risk of ASCVD and all-cause mortality even when accounting for total LE8 score. Future studies should investigate the role of modifying inflammatory burden in addition to LE8 components in prevention of adverse health outcomes.

**Journal Subject Codes:** Cardiovascular disease, Atherosclerosis, Life’s essential 8, Cardiovascular health, aging, inflammation, sleep

## Introduction

Twelve years after the original definition of cardiovascular health (CVH) (1), the American Heart Association (AHA) presented an update of and enhancement to the means for measuring and monitoring CVH (2). The AHA’s “Life’s Essential 8” (LE8) addresses the current context of public health and the challenge of achieving greater health for all (2). Life’s Essential 8 is the updated model to understand, monitor, and modify CVH across the life course, in diverse settings, and for all people. LE8 builds on LE7 by including sleep duration as a newly added metric. Sleep is a multidimensional construct with overlapping components including duration and several measures of sleep quality, however LE8 included only sleep duration (3).

Despite increasing evidence that inflammation is a risk factor for atherosclerotic cardiovascular diseases (ASCVD), LE8 does not include inflammation as a component, (4–6). High circulating levels of interleukin-6 (IL-6), which modulates inflammation, have been identified as a risk factor for adverse ASCVD outcomes and all-cause mortality (7). Additionally, high-sensitive C-reactive protein (hsCRP), can also be used to predict cardiovascular risk (8; 9). With recent data showing that lifestyle and therapeutic interventions alter inflammatory levels, hsCRP and/or IL-6 could be modifiable parameters for ideal cardiovascular health. However, no comprehensive data examine the role of inflammation as an additional factor to LE8.

The objective of the present study was to evaluate LE8 Score in the community-based Heart Strategies Concentrating on Risk Evaluation (Heart SCORE) study and its association with long-term ASCVD events and mortality. Further, we examined the performance of inflammatory markers (measured by IL-6 and hsCRP) on this association.

## Methods

### Study design

Heart SCORE is a community-based study with balanced representation of White and Black US residents in the greater Pennsylvania region (10). Men and women 45-75 years old were recruited through community and faith-based educational and screening programs. All participants underwent a baseline visit between 2003-2005 and annual follow-up to determine traditional and novel cardiovascular risk factors (11), tabulate CVD events, and assess measures of subclinical atherosclerosis (12; 13). The study was approved by the Institutional Review Board at the University of Pittsburgh. All study subjects provided written informed consent.

### Demographic Variables

Age, race, sex, education level and annual income, were obtained by self-report at the baseline visit. Race was self-identified as Black or African American, White, Asian, American Indian or Native Alaskan, Native Hawaiian or Pacific Islander, or Other. Education was categorized as “Less than high school”, “High school”, “Some college” or “College graduate or higher”. Socioeconomic risk was defined as having an annual income of less than $10,000 or an educational attainment less than a high school diploma.

### Past Medical History

At baseline, participants self-reported a history of clinical cardiovascular disease (CVD) - including CHD, heart failure and stroke-the history of CVD risk factors-hypertension, diabetes, hyperlipidemia- and the use of medications for treating hypertension, dyslipidemia or abnormal glucose metabolism.

### Definitions of Life’s Essential 8 (LE8) Score

A modified version of the original AHA LE8 score was developed for this study (**Supplementary Table 1**) (10). Using the eight components including (1) diet, (2) physical activity, (3) tobacco/nicotine exposure, (4) sleep, (5) body mass index, (6) non–high-density lipoprotein (non-HDL) cholesterol, (7) blood glucose, and (8) blood pressure, the total Life’s Essential 8 score was created. Modifications were made to adapt physical activity, diet, and sleep parameters available in the Heart SCORE study. Physical activity was evaluated with the previously used Lipid Research Clinic Questionnaire, which includes questions about the type and frequency of activity during work and leisure time (10; 14). Instead of using the DASH-style eating pattern as proposed by the AHA, we analyzed weekly servings of green vegetables and fruits for dietary assessment, as we used previously (10; 15) For sleep we utilized a preexisting modified version of the PSSQ questionnaire in Heart SCORE study, which is a validated instrument for sleep quality and insomnia assessment (4; 10; 16).

Each health metric was scored from 0 to 100, with a lower score indicating the least healthy while a higher score indicating the healthiest in accordance with work by Lloyd-Jones and colleagues (2)

The mean LE8 score for each individual was derived by summing the eight health metrics and dividing them by 8. The score was examined as both a continuous score as well as quartiles (quarters) of distribution in the analyses.

### Biofluid markers

Biofluid markers were measured with previously demonstrated methods through blood samples at baseline Heart SCORE visit (11; 13). Measurement of lipids was performed with fasting venous blood using standard laboratory techniques at the University of Pittsburgh Medical Center Clinical Laboratories. Serum hsCRP was measured using an immunoturbidimetric assay on the Roche P modular system (Roche Diagnostics, Indianapolis, Indiana). Serum IL-6 was measured using commercially available enzyme-linked immunosorbent assay kits (R&D Systems, Minneapolis, Minnesota). Measurement of fasting blood glucose was performed using the glucose oxidase method.

### Incident Outcomes

The primary outcome was a composite of ASCVD mortality and nonfatal ASCVD events, defined as nonfatal myocardial infarction, acute ischemic syndrome, coronary revascularization (percutaneous coronary intervention or coronary artery bypass graft). All events were tabulated by annual telephonic or in-person follow up, confirmed by hospital record review, and adjudicated by staff cardiologists. Mortality was ascertained and classified as ASCVD death by reviewing death certificates and hospital records. All-cause mortality and all cancer deaths were included as secondary outcomes adjudicated by trained physicians after review of medical records. Outcomes were analyzed until December 31, 2020 with a median of 12.1 (9.2, 12.3) years follow up period.

### Statistical analyses

Descriptive statistics were used to characterize demographics, components of the LE8 score, and inflammatory markers for patients at baseline, summarized overall and by sex. Statistical testing for differences in distributions of study variables by sex used Welch’s t-test for continuous variables and Chi-squared test with Yates’s continuity correction for categorical variables. The association between LE8 and incident ASCVD and mortality events was assessed with Cox regression and presented as hazard ratios and corresponding 95% confidence intervals. Multivariable models adjusted for age, sex, and race. An alpha level of .05 was used for all statistical tests. Statistical analysis was conducted in R version 4.4.2 (17).

## Results

### Demographic and clinical characteristics

A total of 1,869 participants with complete information on LE8 metrics and incident outcomes were included. The baseline characteristics and the distribution of the components of CVH by sex are outlined in **Table 1**. The mean age of the study population is 59±7.5 years at study entry, 41.9% was Black and a majority of individuals had at least some College education with 49.1% that have graduated from college or higher. Only 8% were classified as having socioeconomic risk.

**Table 1.** Baseline characteristics of study population and Life’s Essential 8 metrics by sex. Heart SCORE study, 2003-2020.

| *Characteristic* | *Total*<br>*(n=1869)* | *Men*<br>*(n=642)* | *Women*<br>*(n=1227)* | *P value\** |
| --- | --- | --- | --- | --- |
| **Age, mean (SD)** | 59.0 (7.45) | 59.4 (7.55) | 58.9 (7.39) | .14 |
| **Race, n (%)** |  |  |  | < .01 |
| White | 1037 (55.5) | 383 (59.7) | 654 (53.3) |  |
| Black or African American | 783 (41.9) | 237 (36.9) | 546 (44.5) |  |
| Other | 49 (2.6) | 22 (3.4) | 27 (2.2) |  |
| **Education level, n (%)** |  |  |  | < .01 |
| Less than high school | 41 (2.2) | 20 (3.1) | 21 (1.7) |  |
| High school | 308 (16.5) | 86 (13.4) | 222 (18.1) |  |
| Some college | 602 (32.2) | 167 (26.1) | 435 (35.5) |  |
| College graduate or higher | 916 (49.1) | 368 (57.4) | 548 (44.7) |  |
| **Socioeconomic risk, n (%)** | 136 (8.0) | 47 (7.8) | 89 (8.1) | .96 |
| **Cardiovascular Health Metrics** |  |  |  |  |
| **Diet Score, mean (SD)** | 60.9 (17.7) | 61.1 (18.1) | 60.4 (17.0) | .39 |
| **Physical Activity Score, mean (SD)** | 61.8 (24.0) | 65.6 (23.5) | 59.9 (24.0) | < .001 |
| **Tobacco Score, mean (SD)** | 69.0 (33.1) | 65.9 (33.7) | 70.7 (32.6) | .003 |
| **Sleep Score, mean (SD)** | 54.6 (30.5) | 56.9 (30.4) | 53.4 (30.5) | .02 |
| **BMI, kg/m<sup>2</sup>, mean (SD)** | 30.0 (6.31) | 29.9 (5.65) | 30.1 (6.63) | .47 |
| **Blood Lipids** |  |  |  |  |
| **Non-HDL Cholesterol, mg/dL, mean (SD)** | 156 (39.8) | 151 (37.6) | 158 (40.6) | < .001 |
| **Cholesterol Treatment, n (%)** | 394 (21.1) | 156 (24.4) | 238 (19.4) | .01 |
| **Blood Glucose** |  |  |  |  |
| **Glucose, mg/dL, mean (SD)** | 98.8 (25.7) | 102 (26.3) | 97.2 (25.2) | < .001 |
| **Diabetes, n (%)** | 186 (10.0) | 47 (7.3) | 93 (7.6) | .99 |
| **Glucose Treatment, n (%)** | 140 (7.5) | 47 (7.3) | 93 (7.6) | .92 |
| **Blood Pressure** |  |  |  |  |
| **Systolic Blood Pressure, mmHg, mean (SD)** | 136 (19.5) | 138 (18.4) | 135 (20.0) | .004 |
| **Diastolic Blood Pressure, mmHg, mean (SD)** | 80.8 (10.3) | 82.4 (10.4) | 80.0 (10.1) | < .001 |
| **Blood Pressure Treatment, n (%)** | 779 (41.7) | 259 (40.5) | 520 (42.4) | .45 |
| **Total Life Essential 8 Score, mean (SD)** | 60.7 (11.3) | 60.5 (10.9) | 60.7 (11.5) | .70 |
| **Inflammation** |  |  |  |  |
| **IL-6, pg/mL, mean (SD)** | 2.20 (2.02) | 2.07 (1.74) | 2.27 (2.15) | .03 |
| **hsCRP, mg/dL, mean (SD)** | 3.16 (6.39) | 2.50 (6.13) | 3.50 (6.50) | .001 |

**\*Calculated using Welch's t-test for continuous variables and Chi-square test for categorical variables.**

### Life’s Essential 8 Score

The distribution of participants according to categories of Life’s Essential 8 Score metrics is available in **Supplementary Table 2**. The mean LE8 Score was 60.7 ±11.3 points. Quartiles cut-points for the total LE8 Score were 53.1, 60.6, and 68.8 [25^th^, 50^th^, 75^th^ IQR]. **Figure 1** displays the distribution of the participants of the Heart SCORE study according to Life’s Essential 8 Score.

**Figure 1.**
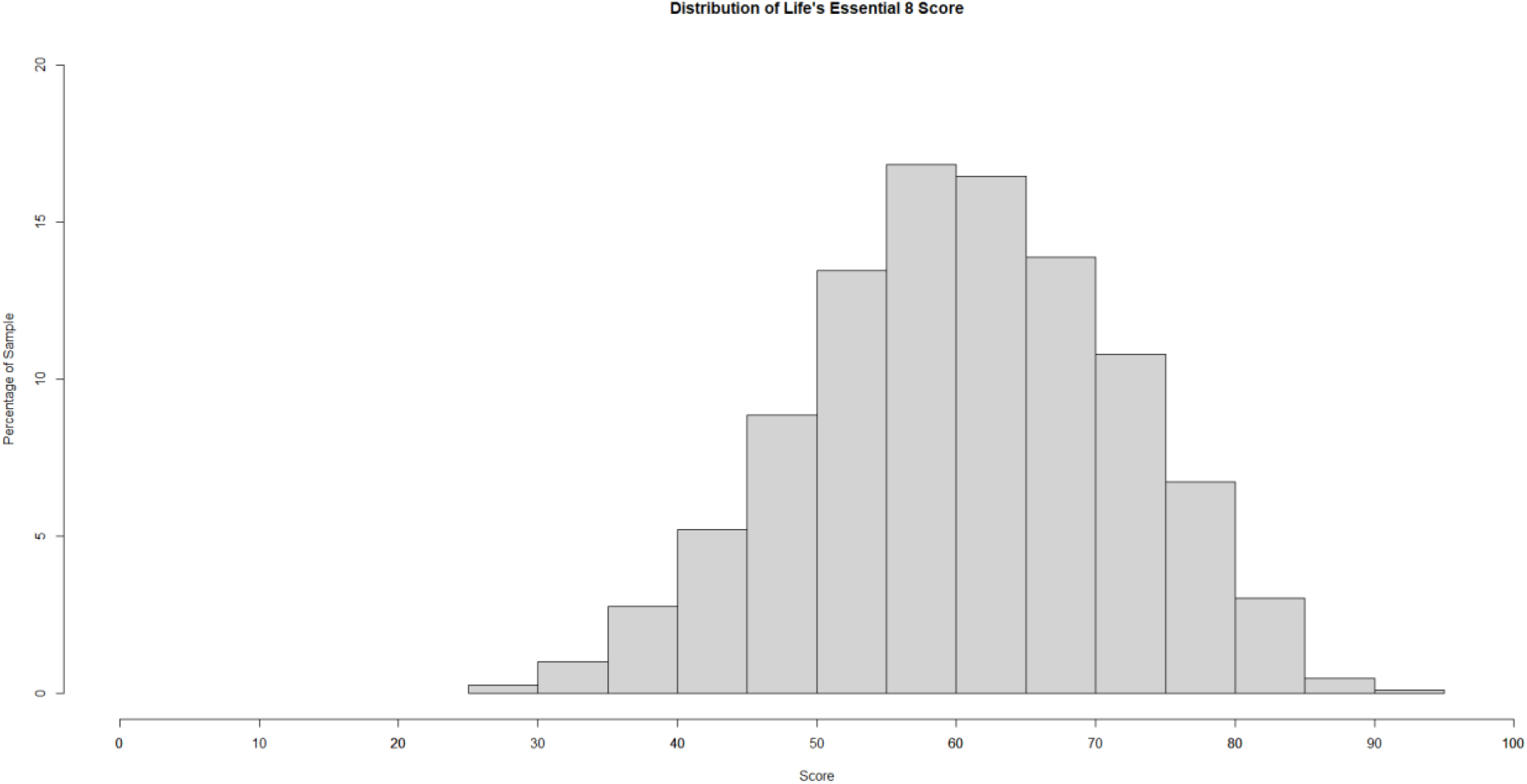
Distribution of participants according to Total Life’s Essential 8 Score, HEART SCORE study, 2003-2021.

### LE8 with ASCVD and All-Cause Mortality

**Figures 2 to 4** display survival curves for all-cause mortality, ASCVD and cancer deaths, respectively, among participants in the highest versus lower Quartile of Life’s Essential 8 Score from 2003 to 2020. All models were adjusted by age, sex, race and socioeconomic risk.

**Figure 2:**
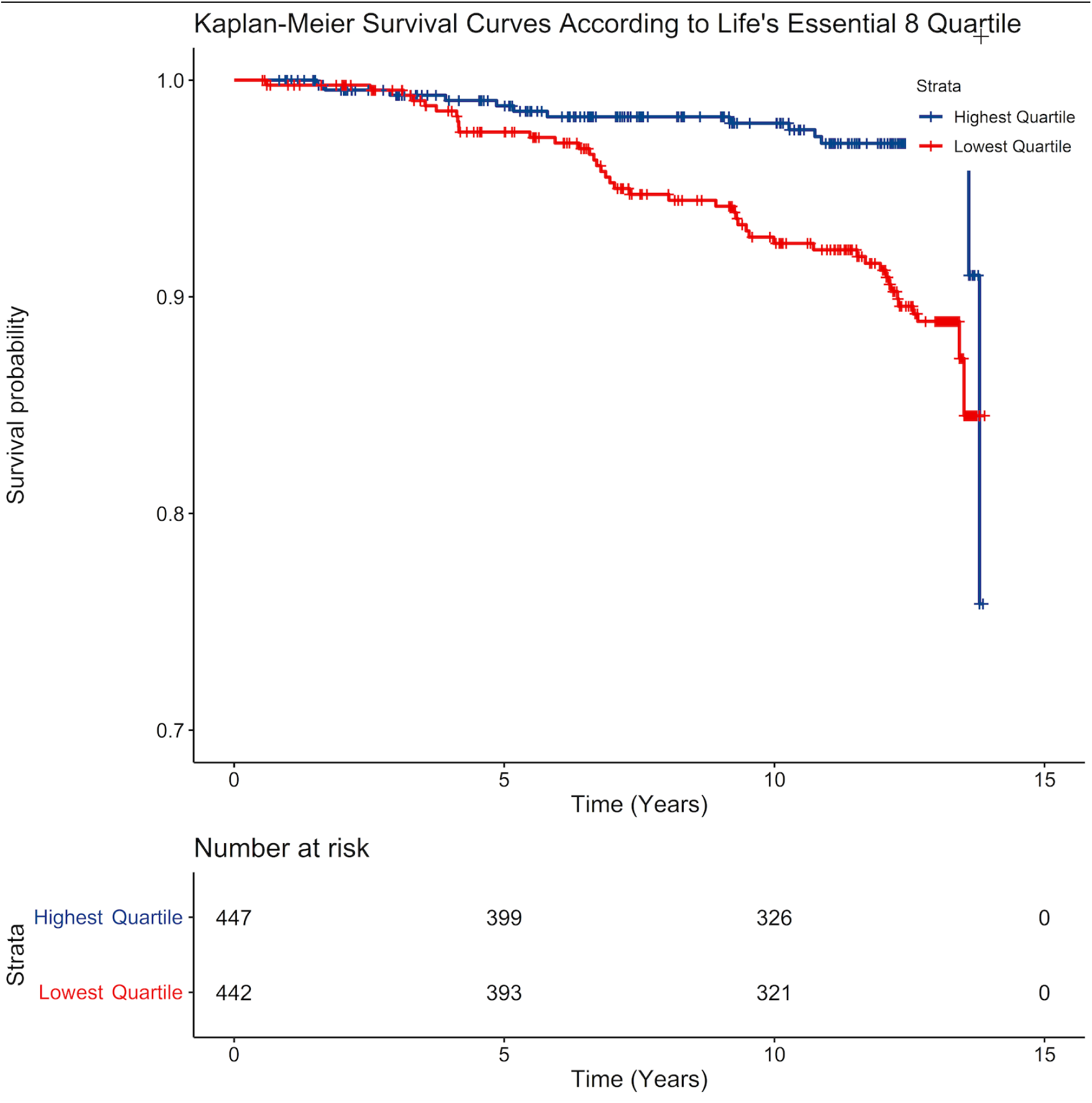
All-cause mortality among participants in the highest versus lowest Quartile of Life’s Essential 8 Score. HEART Score Study, 2003-2020.

**Figure 3:**
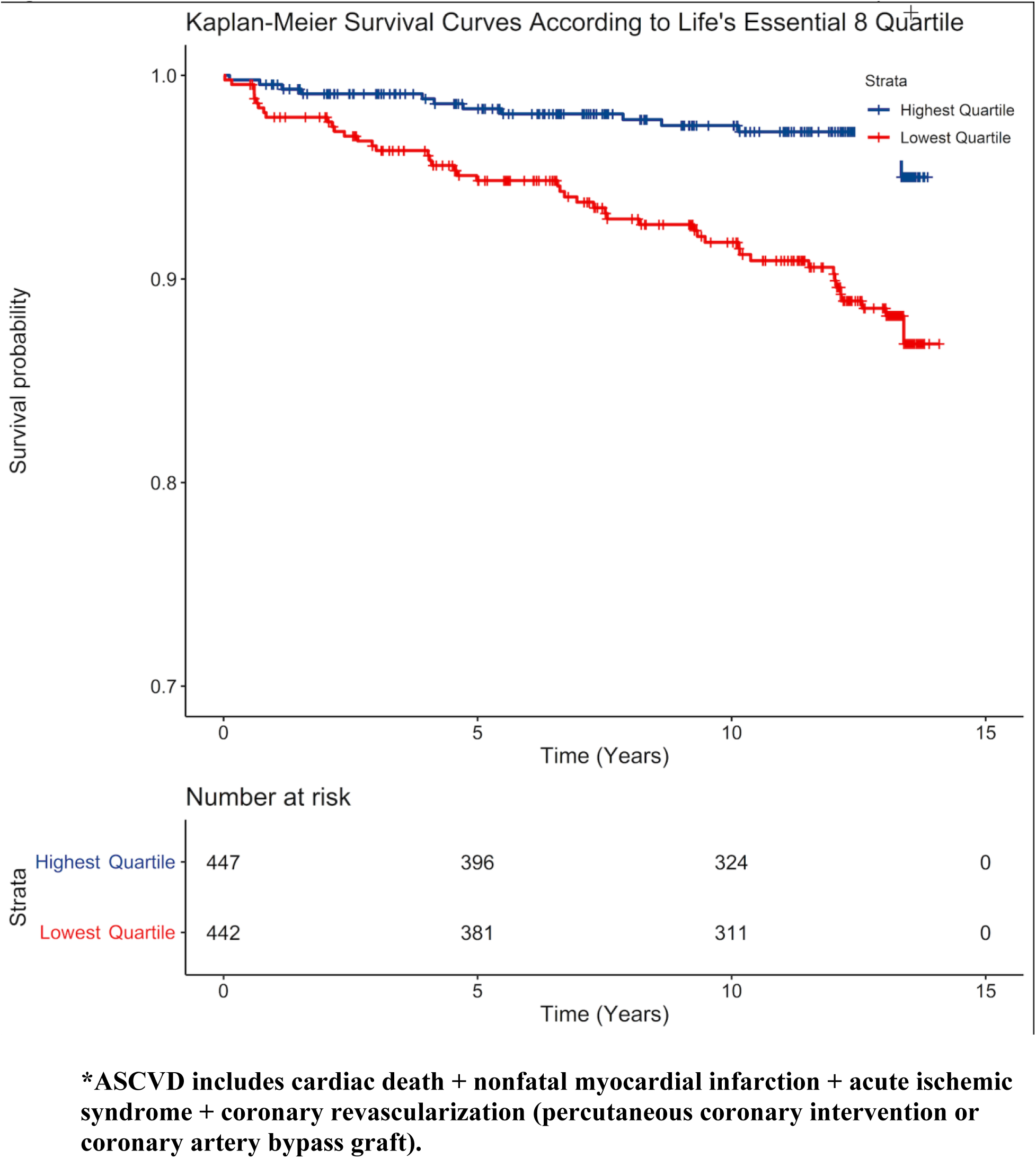
Atherosclerotic Cardiovascular Disease (ASCVD*) among participants in the highest versus lowest Quartile of Life’s Essential 8 Score. HEART Score Study, 2003-2020.

**Figure 4:**
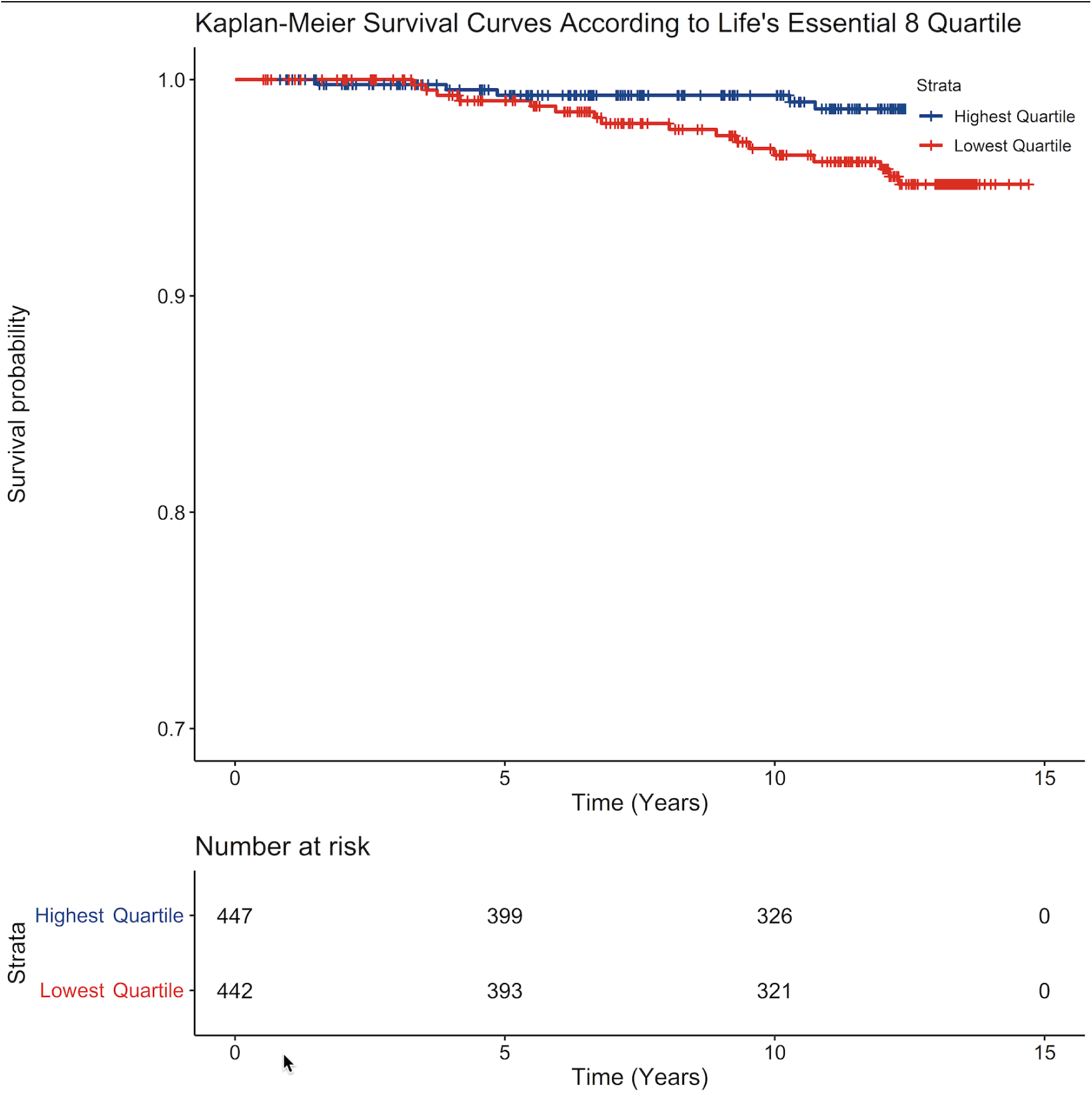
Cancer deaths among participants in the highest versus lowest Quartile of Life’s Essential 8 Score. HEART Score Study, 2003-2020.

Compared to lowest quartile, all higher quartiles of LE8 Score were significantly associated with lower risk of all-cause mortality and ASCVD over a median of 12 years of follow-up. In both cases, a dose-response effect was observed exhibiting a protective effect of LE8 Score over these outcomes (**Table 2)**.

**Table 2.** Association between Life’s Essential 8 Scores and health outcomes. Multivariable survival analysis at Heart SCORE study, 2003-2020

| Covariates | All-cause mortality | ASCVD | Cancer death |
| --- | --- | --- | --- |
|  | HR (95% CI) | HR (95% CI) | HR (95% CI) |
| Total LE8 Score |  |  |  |
| Highest quartile (vs lowest) | 0.38 (0.21-0.69)* | 0.26 (0.14-0.51)* | 0.45 (0.18-1.14)<sup>ns</sup> |
| Second highest quartile (vs lowest) | 0.45 (0.26-0.76)** | 0.51 (0.30-0.85)*** | 0.62 (0.27-1.39)<sup>ns</sup> |
| Second lowest quartile (vs lowest) | 0.59 (0.36-0.98)** | 0.30 (0.16-0.55)* | 0.90 (0.44-1.86)<sup>ns</sup> |
| Age | 1.07 (1.04-1.10)* | 1.06 (1.03-1.09)* | 1.07 (1.03-1.12)** |
| Female Sex | 0.41 (0.28-0.60)* | 0.43 (0.28-0.65)* | 0.46 (0.26-0.81)** |
| Black Race | 1.33 (0.90-1.98)<sup>ns</sup> | 1.10 (0.71-1.70)<sup>ns</sup> | 1.11 (0.61-1.99)<sup>ns</sup> |
| Socioeconomic Risk | 1.44 (0.79-2.61)<sup>ns</sup> | 1.25 (0.62-2.52)<sup>ns</sup> | 2.64 (1.27-5.46)** |

\*p-value <0,001
\*\*p-value <0,05
\*\*\*p-value=0,01
ns= p-value >=0,05
**ASCVD**= Atherosclerotic cardiovascular diseases (cardiac death + nonfatal events: myocardial infarction, acute ischemic syndrome, percutaneous coronary intervention or coronary artery bypass graft).

In terms of individual LE8 metrics, ideal level of blood glucose was associated with significantly lower risks of ASCVD (HR 0.26 [0.12-0.58], p=0.001) and a similar trend was seen for ideal blood pressure (HR 0.45 [0.18-1.12], p=0.086). For all-cause mortality, no smoking emerged as the main protective factor after adjusting for all other LE8 metrics as wells as age, sex, race and socioeconomic risk (HR 0.29 [0.16-0.53], p<0.001). Ideal blood glucose and ideal blood pressure showed similar trends (HR 0.47 [0.22-1.0], p=0.051 and HR 0.42 [0.18-1.01], p =0,052, respectively). No significant associations were noted with sleep quality (**Table 3).**

**Table 3.**
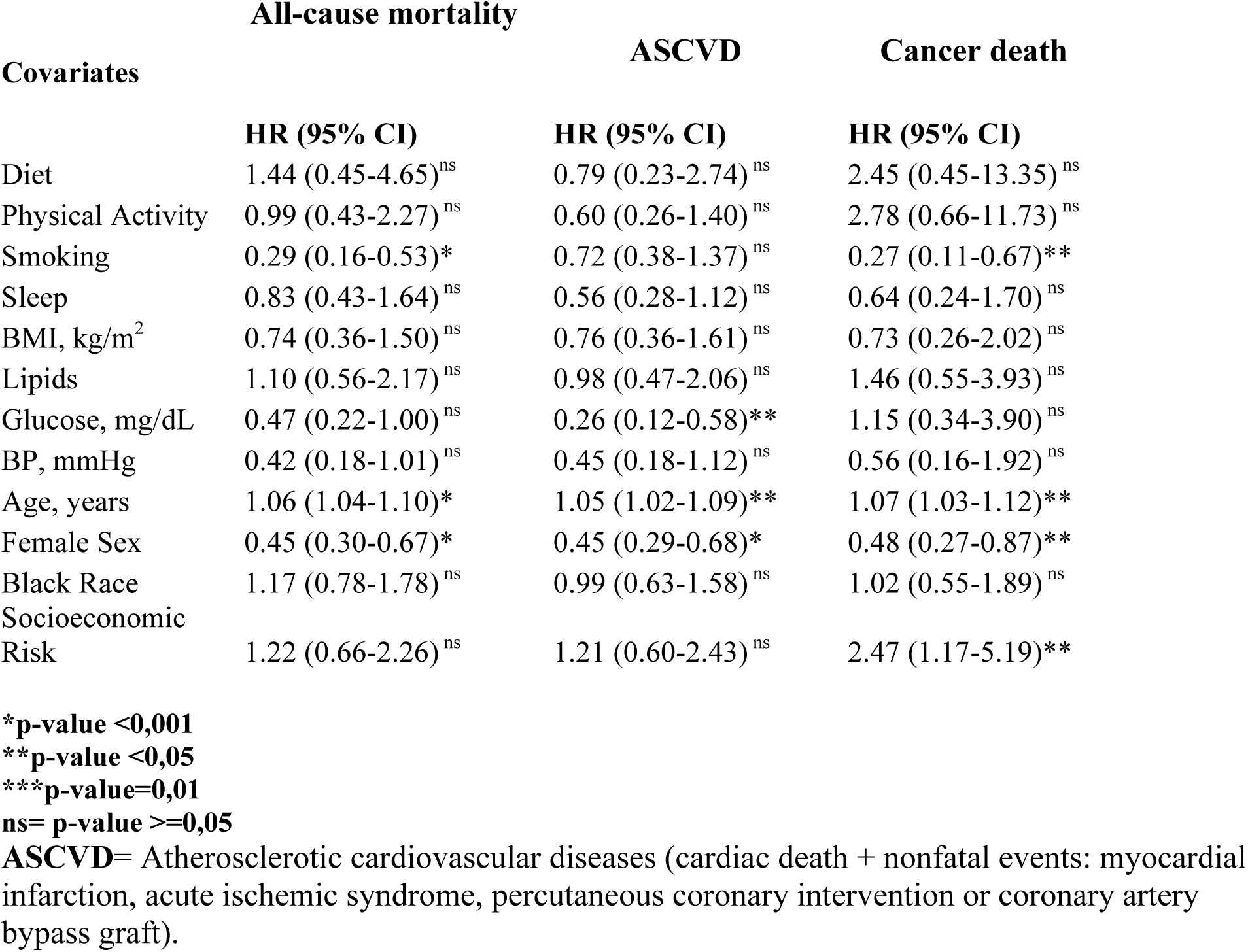
Association between individual Life’s Essential 8 metrics and health outcomes. Multivariable survival analysis at Heart SCORE study 2003-2020

### LE8 and Cancer Death

As displayed in **Table 2**, multivariable survival models showed no significant association between LE8 Score and cancer deaths, however, a protective trend was observed when comparing highest versus lowest quartile of LE8 Score (HR 0.45 [0.18-1.14], p=0.093). Also, when individual LE8 metrics were included in multivariable survival models, no smoking was associated with 73% lesser risk of cancer death compared to current smoking (HR 0.27 [0.11-0.67], p=0.004).

### Impact of Inflammatory Markers

Continuous multivariable survival models including LE8 score and inflammatory markers (IL-6 and hsCRP) showed that IL-6 was independently associated with higher risk of ASCVD mortality and non-fatal ASCVD events (HR 1.54 [1.05-2.25], p=0.028) but not cancer or all-cause mortality, while hsCRP showed no association with any outcome (**Table 4**). In further analysis with IL-6 broken down into quartiles, the lowest quartile of IL-6 compared to the highest had a lower risk of all-cause mortality (HR 0.52 [0.30-0.90], p=0.019) but not ASCVD or cancer death. In fully adjusted models, we did not find a significant association of hsCRP with ASCVD, cancer, or all-cause mortality (**Supplemental table 3)**.

**Table 4.** Association between Life’s Essential 8 Score and IL-6 and health outcomes. Multivariable survival analysis at Heart SCORE study 2003-2020

| **Covariates** | **All-cause mortality** | **ASCVD** | **Cancer death** |
| --- | --- | --- | --- |
|  | **HR (95% CI)** | **HR (95% CI)** | **HR (95% CI)** |
| Simple 8 Score, Highest Quartile (vs Lowest) | 0.52 (0.30-0.90)** | 0.28 (0.15-0.54)* | 1.05 (0.43-2.58)<sup>ns</sup> |
| Simple 8 Score, Second Highest Quartile (vs Lowest) | 0.81 (0.50-1.31)<sup>ns</sup> | 0.50 (0.30-0.85)** | 1.98 (0.96-4.07)<sup>ns</sup> |
| Simple 8 Score, Second Lowest Quartile (vs Lowest) | 0.58 (0.36-0.94)** | 0.29 (0.15-0.54)* | 1.44 (0.68-3.05)<sup>ns</sup> |
| Age | 1.06 (1.03-1.09)* | 1.06 (1.03-1.10)* | 1.00 (0.96-1.04)<sup>ns</sup> |
| Female Sex | 0.49 (0.34-0.70)* | 0.51 (0.33-0.77)** | 0.86 (0.49-1.48)<sup>ns</sup> |
| Black Race | 1.51 (1.04-2.20)** | 0.84 (0.53-1.31)<sup>ns</sup> | 1.30 (0.73-2.32)<sup>ns</sup> |
| IL-6, Lowest Quartile (vs Highest) | 0.52 (0.30-0.90)** | 0.65 (0.33-1.28)<sup>ns</sup> | 0.78 (0.34-1.81)<sup>ns</sup> |
| IL-6, Second Lowest Quartile (vs Highest) | 0.60 (0.37-0.99)** | 0.91 (0.51-1.62)<sup>ns</sup> | 0.60 (0.29-1.27)<sup>ns</sup> |
| IL-6, Second Highest Quartile (vs Highest) | 0.90 (0.57-1.43)<sup>ns</sup> | 0.98 (0.56-1.69)<sup>ns</sup> | 0.71 (0.35-1.43)<sup>ns</sup> |

**\*p-value <0,001**
**\*\*p-value <0,05**
**\*\*\*p-value=0,01**

## Discussion

Our results show that over a median follow-up of 12 years, lower Life Essential 8 (LE8) score is associated with higher incidence of all-cause mortality and ASCVD mortality and non-fatal ASCVD events in a community-based population with a comparable representation of Black and White individuals as well as men and women. High inflammatory burden, measured by IL-6 levels, was independently associated with adverse health outcomes despite a high LE8 score. Sleep quality was not independently associated with adverse outcomes.

Our findings are unique in exploring the effect of inflammation in the context of LE8 on longitudinal outcomes. To our knowledge, no longitudinal studies have directly examined the residual effect of inflammatory burden (using IL-6 and hsCRP) on health outcomes within the context of LE8. Although recent reports (18; 19) suggest that LE8 and isolated hsCRP may have an inverse relationship, these data lack longitudinal time points. Our results corroborate recent randomized control trial data that suggest that IL-6 but not hsCRP may be associated with higher risk of major adverse cardiovascular events (20; 21). We also found that lower IL-6 levels were associated with lower all-cause mortality across all categories of LE8. Previous research has identified an association with high circulating IL-6 and increased cardiovascular mortality and all-cause mortality (7).

Our work adds to previous reports that have shown positive associations between ideal cardiovascular health and all-cause mortality and cardiovascular diseases (22). The trend between better cardiovascular health and lower cancer deaths that we observed is particularly important given that cancer has become second only to cardiovascular disease for the number of deaths, years of life lost, and DALYs globally in 2019, showing that the global burden of cancer is substantial and growing and that its prevention is a matter of great interest globally ((23)). In our study, the low number of cancer deaths (n=65) probably explain why our results did not reach statistical significance. However, the observed trend is important both clinically and epidemiologically, considering that cancer has become an important cause of death even in presence of cardiovascular diseases, overcoming the classic competitive effect that has been described for cardiovascular diseases and cancer. The fact that cancer and CVD share lifestyle risk factors and that the exposure to such risk factors is occurring at increasingly earlier ages can explain at least in part the new onset cancers in middle age populations (24). In our study cancer death as opposed to cancer incidence was used because cancer incidence data was not available.

Our results highlight that cigarette smoking plays an important role in terms of all-cause mortality, cardiovascular disease and cancer deaths. It is well known that tobacco exposure increases the risk of cardiovascular disease by 2 to 4 times and that has a causal relationship with at least 15 different cancer types of cancer (25). Also, evidence supports that even occasional or social, nondaily basis smoking increases the risk of cardiovascular diseases and cancer, as well as the risk of dying prematurely compared to no smoking (26; 27). On the other hand, smoking cessation is associated with a 50% reduction in coronary heart disease after one year and with a 50% reduction in the risk of cancers of the mouth, throat, esophagus, and bladder after 5 years (28). As demonstrated by Yoo et al., smoking cessation and, to a lesser extent, smoking reduction decreased the risks of cancer, and smoking resumption increased cancer risks in comparison with sustained quitting (29). Thus, tobacco control and smoking cessation remain important public health goals.

Interestingly, our study did not identify a relationship between sleep quality or hsCRP with ASCVD, all-cause mortality, or cancer deaths. One of the foundational studies that led to the inclusion of sleep in the AHA’s updated guidelines evaluated sleep as a measure of cardiovascular health by including it as an additional component of AHA’s previous cardiovascular health guidelines, Life’s Simple 7, rather than as an individual predictor (30).. Studies that examined associations between short sleep duration and coronary artery disease (CAD) show conflicting results (31; 32). Individual components of LE8 occur in conjunction with and may interact with the other factors, so it is not surprising that sleep and other individual components were not associated with adverse outcomes. The lack of relationship of hsCRP with ASCVD that we found may be due to the variation of hsCRP levels and its role as a risk marker rather that a risk factor for adverse health outcomes (33). Though studies have identified hsCRP as a predictor of adverse cardiovascular events (8; 9), Saeed et. al., have showed that hsCRP is not a robust risk factor of ASCVD once other factors are accounted for (34).

There currently is not a consensus for screening or monitoring IL-6 for ASCVD risk stratification and few therapies are available for targeted reduction. Our study raises into question the potential of routine monitoring IL-6 levels as a more robust inflammatory burden marker. Studies have explored anti-inflammatory therapies to reduce adverse cardiac events. CANTOS (Canakinumab Antiinflammatory Thrombosis Study) describes a reduction in adverse cardiac events and a reduction in IL-6 and hsCRP with Canakinumab, a monoclonal antibody targeting interleukin-1b (35). Although there are conflicting data as to whether colchicine reduces the risk of ischemic events (20; 21; 36), individuals who have higher inflammatory burden may benefit from more aggressive risk factor management such as statin therapy initiation or intensification (37). Our results shed light on the importance of continued studies to target different inflammation pathways to reduce adverse health events.

## Limitations

Our study has several limitations. We identify four potential sources of residual confounding in this analytical design: categorization of socioeconomic status variables, measurement error in socioeconomic indicators, use of aggregated socioeconomic status measures, and incommensurate socioeconomic indicators (38). Also, our population is subject of referral and healthy volunteer biases, which can be recognized by a low prevalence of smoking (10.6%) in comparison with general population of the same age (28) and an overall high level of education. Although this can limit the generalizability of our findings, the bias is toward a conservative scenario, and the prevalence of healthy behaviors and factors in the real world could be much lower. Additionally, as in other epidemiologic studies, misclassification could occur especially when nutritional and physical activities were evaluated. The PrimeScreen questionnaire was designed to evaluate quality of food more than quantity, so the total daily consumption of fruits and vegetables could not be derived directly from that source and what we used here approximates the recommended 5 servings per day. Furthermore, the other nutritional components recommended by AHA (fish, fiber-rich whole grains, sodium and sugar-sweetened beverages) were not assessed in the current investigation. Sleep duration was the latest factor added to LE8, but we used data from a sleep quality based on an insomnia PSSQ-I questionnaire due to availability. However, we used the which evaluates quality of sleep and insomnia symptoms which have been linked with ASCVD risk (39). Finally, we assess biofluid markers at one time point and as such; markers including hsCRP and lipid levels are subject to change over short and longitudnal time points.

Our study’s strengths include that the longitudinal results here reported correspond to the analysis of data from a community-based participatory research study, in which direct involvement of the community, community-based organizations and faith-related institutions occurred in the recruitment process and constitutes now a source of information to study diverse communities over time.

In conclusion, our findings indicate that better levels of LE8 Score and metrics are associated with lower risk of all-cause mortality and ASCVD in the Heart SCORE study population. Additionally, our results show a relationship between higher levels of IL-6 and increased ASCVD and all-cause mortality. These results underscore the role of improving and monitoring cardiovascular health as a mean to reduce the burden of mortality and morbidity among diverse communities. Further studies are needed to confirm the role of IL-6 and other inflammation markers as additional factors to LE8.

## Data Availability

The data that support the findings of this study are available from the corresponding author upon reasonable request.

